# Menstrual hygiene management among reproductive-aged women with disabilities in Bangladesh

**DOI:** 10.1101/2024.05.23.24307772

**Authors:** Md Nuruzzaman Khan, Shimlin Jahan Khanam, Atika Rahman Chowdhury, Rashed Hossain, Md Awal Kabir, Md Badsha Alam

## Abstract

**Background:** Women with disabilities in low- and middle-income countries face unique challenges in managing menstruation, affecting their health, dignity, and quality of life.

**Objective:** This study explored menstrual hygiene management (MHM) practices among reproductive-aged women with disabilities in Bangladesh and its impact on social participation.

**Methods:** We analyzed data from 51,535 women from the 2019 Multiple Indicator Cluster Survey (MICS). The outcome variables were: (i) material used to manage blood flow while menstruating (appropriate, inappropriate), (ii) availability of a private place for washing and changing menstruation rags (yes, no), and (iii) impacted in attendance of social activities, school, or work during menstruation (yes, no). Disability status was considered as major explanatory variable and categorized as no disabilities, moderate disabilities, and severe disabilities. Associations of outcome variables with explanatory variable were determined using a multilevel multinomial logistic regression model adjusted for individual-level factors, household-level factors, and community-level factors.

**Results:** Among the sample, 2.7% reported severe disability and 19.2% moderate disabilities, with vision-related disabilities (12.2%) being the most prevalent, followed by cognitive (9.5%) and walking disabilities (8.2%). Overall, 4% reported using inappropriate materials for menstrual flow, rising to 8.0% among those with severe disabilities and 5.8% for moderate disabilities. Women with moderate to severe disabilities had 33-47% lower odds of using appropriate materials and 34-44% lower odds of having a private place to change at home. Additionally, women with severe disabilities were 1.66 times more likely to report that menstruation impacted their social activities.

**Conclusion:** Women with disabilities in Bangladesh face significant barriers to effective MHM. Addressing these issues requires targeted interventions, including disability-inclusive menstrual health education, improved access to menstrual products and sanitation facilities, and community initiatives to reduce stigma.

## Background

Management of menstrual health and hygiene is essential for women’s well-being and empowerment. However, it remains an unmet public health need globally, particularly in low- and middle-income countries (LMICs), where approximately 500 million women and girls lack adequate facilities for menstrual management [1–3]. This shortfall is often compounded by a lack of access to menstrual hygiene products [1]. While one-third of women worldwide lack access to basic sanitation facilities, this figure is significantly higher in LMICs, although accurate estimates are lacking [4, 5]. Moreover, cultural taboos and social stigmas surrounding menstruation exacerbate the challenges faced by women and girls in effectively managing their menstrual hygiene [6–8]. Consequently, inadequate menstrual hygiene management (MHM) contributes to increased health risks, including urinary tract infections, secondary infertility, anxiety, depression, and decreased self-esteem [9, 10].

Persons with disabilities constitute one of the largest minority groups worldwide, with approximately 15% of the global population [11, 12]. Around 80% of them live in LMICs, highlighting the disproportionate impact on these regions [13, 14]. This demographic encompasses various impairments, including physical, sensory, intellectual, and mental health disabilities. Furthermore, there is a global trend of increasing disability prevalence with particularly more pronounced in LMICs, attributed partly to population aging, the rising incidence of chronic diseases, and existing burden of higher poverty [15–20]. The advancement of medical technology also contributes to this trend by enabling the survival of individuals who would otherwise not have survived [21].

Persons with disabilities in LMICs often report poorer health outcomes, particularly due to limited access to health-related information [18, 22]. Mobility restrictions and lower participation in education further contribute to these disparities [23, 24]. Additionally, they often rely on social support networks or family members for their basic needs, including healthcare [25–27]. This dependence creates challenges in prioritizing and addressing health issues due to limited options

[28, 29]. Mobility restrictions further exacerbate this situation by hindering access to freely available services, typically accessible to the general population [23].

These structural issues often result in MHM for persons with disabilities being overlooked in LMICs [30]. Existing studies in this setting often focus on the general population and their MHM practices, with findings revealing concerning scenarios, such as the reuse of the same old cloth for several months [31–33]. Commonly reported barriers to safe MHM for persons with disabilities include cultural beliefs, family environment, education level, poverty, lack of appropriate sanitation facilities, cost, and access to menstrual hygiene products [32, 33]. These factors, combined with the structural barriers mentioned above, suggest very poor MHM among persons with disabilities [32–35]. Bangladesh exemplifies this experience, although relevant studies conducted so far are lacking, with a few studies focusing on specific disability types or small areas [36–40]. Therefore, this study aims to explore MHM practices among reproductive-aged women with disabilities in Bangladesh.

## Methods

### Study setting and sampling techniques

We extracted data from the 2019 Multiple Indicator Cluster Survey (MICS) and analyzed. This nationally representative household-based cross-sectional survey was conducted by the Bangladesh Bureau of Statistics (BBS) as part of UNICEF’s initiative to collect data on the well-being of mothers and children in LMICs through standardized surveys. The survey took place from January 19 to June 2019 and sampled households using a two-stage stratified cluster sampling technique, covering all 64 districts of Bangladesh. Initially, 1,300 Primary Sampling Units (PSUs), areas with an approximate population of 200 to 300 households, were selected based on the 2011 Bangladesh Population and Housing Census. In the second stage of sampling, 20 households were chosen from each PSU using systematic random sampling techniques, resulting in a total sample of 64,400 households. Of these, 61,602 households were occupied, and 61,242 were successfully interviewed, yielding a household response rate of 99.4%. Among the selected households, 68,711 eligible women met the inclusion criteria: reproductive-aged (15-49 years) married women who either resided in the selected households as usual residents or stayed there during the most recent night. Of these eligible women, 64,378 were successfully interviewed, resulting in a response rate of 93.7%. Detailed information about the methodology and sampling structure employed in the survey has been published in the respective survey report [41].

### Analyzed sample

Data from 51,535 women from the original sample who met the inclusion criteria for this study were analyzed. The inclusion criteria were as follows: (i) reproductive-aged women (15-49 years), (ii) availability of information on women who menstruated in the last 12 months and reported related information, (iii) women who reported not being able to participate in social activities, school, or work during their last menstruation in the last 12 months, and (iv) women who reported their disability status.

### Outcome variables

We considered three aspects of MHM and related consequences as outcome variables. These included: (i) material used to manage blood flow while menstruating (appropriate, inappropriate), (ii) availability of a private place for washing and changing menstruation rags (yes, no), and (iii) impacted in attendance of social activities, school, or work during menstruation (yes, no).

Relevant information on the use of appropriate materials during the last menstruation was collected by asking women two specific questions: “Did you use any materials such as sanitary pads, tampons, or cloth?” Women who responded “Yes” to this question were asked, “What type of materials did you use in your last menstruation?” The response options were (i) sanitary napkin, (ii) cotton pad, (iii) tissue, (iv) cloth, and other. The usage of appropriate materials during the last menstruation was categorized for those who reported using sanitary pads and new cloths, otherwise categorized as inappropriate material used. For the availability of a private place for washing and changing at home during menstruation, women were asked the question, “During your last menstrual period, were you able to wash and change in privacy while at home?” The response options were “Yes” and “No”. Furthermore, we generated a separate variable for whether menstruation impacted in participation of social activities, including schooling and work. During survey women were asked “Due to your last menstruation, were there any social activities, school, or workdays that you did not attend?” The response options were “Yes” and “No”. We reclassified the responses as “impacted” if women responded yes and otherwise classified as “not impacted”

### Explanatory variables

The level of disability (no disability, moderate disability, and severe disability), including the types of disability, was the primary exposure variable for this study. These variables were assessed using a set of six questions from the Washington Group Short Set of Disability based on the World Health Organization’s International Classification of Functioning, Disability and Health [42]. The six questions encompassed all six types for assessing disability: vision, hearing, mobility, self-care, communication, and cognition-related disability. The questions were: (1) Do you experience difficulty with your vision, even while using glasses? (2) Do you experience difficulty with your hearing, even with the assistance of a hearing aid? (3) Do you encounter difficulty with walking or ascending stairs? (4) Do you face challenges with remembering or maintaining focus? (5) Do you find self-care activities, such as bathing, dressing, feeding, toileting, etc., to be problematic? and (6) Do you struggle with communication, such as comprehending or being understood? The possible responses were four options for each question to respond: (a) no difficulty, (b) some difficulty, (c) a lot of difficulty, and (d) cannot do or unable to see/hear/walk/remember/self-care/communicate at all. We categorized the disability to analyze it in two different ways. Firstly, we generated a variable with three mutually exclusive categories: (i) no disability, (ii) moderate disability, and (iii) severe disability. Women were classified as “no disability” if they responded ‘no difficulty at all’ to all six questions, “moderate disability” if women reported ‘some difficulty’ to at least one item, and “severe disability” if they reported ‘a lot of difficulty or cannot function at all’ to at least one item. Additionally, we included specific variables for each type of disability individually. For each question, we generated a binary variable––“yes” if women reported having ‘some difficulty and a lot or cannot do at all’ and “no” if they reported having ‘no difficulty’. Furthermore, the types of disabilities were also considered as explanatory variables.

### Covariates

This study incorporated several covariates to assess the association between the exposure variables and outcome variables. We selected covariates through a comprehensive process. Initially, we conducted a comprehensive search of previous literature to identify relevant variables using several databases based on LMICs and Bangladesh [2, 8–10, 26, 29, 30, 32, 35–37, 39, 43]. Next, we ensured their availability in the MICS dataset. Then we assessed them for their statistical associations with the outcome variables. Finally, variables that were statistically significant were included in the analysis and categorized into three levels: individual-level factors, household-level factors, and community-level factors. Individual-level factors included women’s age (18-24 years, 25-34 years, and ≥35 years), education (pre-primary, primary, secondary, and higher), exposure to mass media (not exposed, exposed). Household wealth quintile (poorest, second, middle, fourth, and richest) was included as the household-level factor. The community-level factors included were place of residence (urban and rural), and administrative divisions (Barishal, Chattogram, Dhaka, Khulna, Mymensingh, Rajshahi, Rangpur, Sylhet).

### Statistical analysis

Descriptive statistics were used to summarize the characteristics of the respondents. The surveys we analyzed had a nested structure, with individuals nested within households and households nested within clusters. Previous studies have shown that using simple logistic regression models on hierarchical data produces less precise results. Therefore, we determined the associations of outcome variables with explanatory variables using a multilevel mixed-effects binary logistic regression model. We ran both unadjusted and adjusted models separately for each outcome. Unadjusted models included only outcome and exposure variables, while adjusted models accounted for individual, household, and community-level covariates. Sampling weight and complex survey design were considered in all analyses. Results were presented as unadjusted (cOR) and adjusted odds ratios (aOR) with their corresponding 95% confidence intervals (95% CI). This study was designed and reported following the Strengthening the Reporting of Observational Studies in Epidemiology (STROBE) guidelines. All statistical analyses were performed using Stata software (version 17.0).

## Results

### Background characteristics of the respondents

Table 1 presents background characteristics of the respondents. The mean age of the respondents was 30.8 (±8.5) years. Around 42% of the women had secondary level education. Nearly 70% of the total respondents reported no exposure to mass media. Approximately, 18% of women belonged to poorest household. Geographically, a significant portion, comprising almost 76% women resided in urban areas. Moreover, 26% of the analyzed women were from Dhaka, followed by Chattogram (19.3%) as their administrative region.

**Table 1:** Background characteristics of the respondents, MICS, 2019 (N= 51,535)

| Characteristics | N | Percent | 95% CI |
| --- | --- | --- | --- |
| **Individual level factors** |  |  |  |
| **Ages of women, mean (±SD)** |  | **30.8 (8.5)** |  |
| ≤24 years | 14,730 | 28.6 | 28.1-29.0 |
| 25-34 years | 19,906 | 36.7 | 36.2-37.2 |
| ≥35 years | 17,898 | 34.7 | 34.3-35.2 |
| **Women's education** |  |  |  |
| Pre-primary | 8,123 | 15.8 | 15.3-16.2 |
| Primary | 12,312 | 23.9 | 23.4-24.4 |
| Secondary | 21,416 | 41.6 | 41.0-42.1 |
| Higher | 9,683 | 18.8 | 18.2-19.4 |
| **Exposure to mass media** |  |  |  |
| Not exposed | 15,519 | 30.1 | 29.4-30.9 |
| Exposed | 36,015 | 69.9 | 69.1-70.7 |
| **Household level factors** |  |  |  |
| **Wealth index** |  |  |  |
| Poorest | 9,007 | 17.5 | 16.8-18.2 |
| Second | 9,602 | 18.6 | 18.1-19.2 |
| Middle | 10,271 | 19.9 | 19.4-20.5 |
| Fourth | 10,947 | 21.2 | 20.6-21.9 |
| Richest | 11,709 | 22.7 | 21.9-23.5 |
| **Community level factors** |  |  |  |
| **Place of residence** |  |  |  |
| Urban | 12,341 | 24.0 | 23.4-24.5 |
| Rural | 39,194 | 76.0 | 75.5-76.6 |
| **Divisions** |  |  |  |
| Barishal | 2,754 | 5.3 | 5.2-5.5 |
| Chattogram | 9,941 | 19.3 | 18.9-19.7 |
| Dhaka | 13,346 | 25.9 | 25.4-26.4 |
| Khulna | 5,938 | 11.5 | 11.2-11.8 |
| Mymensingh | 3,489 | 6.8 | 6.5-7.0 |
| Rajshahi | 6,736 | 13.1 | 12.8-13.4 |
| Rangpur | 5,652 | 11.0 | 10.7-11.3 |
| Sylhet | 3,678 | 7.1 | 6.7-7.7 |

### Distribution of disability and its types

The percentage distribution of disability levels and types is presented in Table 2. Severe disability was reported by 2.7% of the total sample, followed by 19.2% reporting moderate disability. Among specific domains of disability, vision-related disabilities were found in 12.2% of the total sample, followed by cognitive disabilities (9.5%) and walking disabilities (8.2%).

**Table 2:** Prevalence of types of disability in Bangladesh, MICS, 2019.

| Disabilities level | N | Percent | 95% CI |
| --- | --- | --- | --- |
| No disabilities | 40,284 | 78.2 | 77.7-78.6 |
| Moderate disabilities | 9,876 | 19.2 | 18.7-19.6 |
| Severe disabilities | 1,375 | 2.7 | 2.5-2.8 |
| **Types of disabilities** |  |  |  |
| **Vision related disabilities** |  |  |  |
| No | 45,233 | 87.8 | 87.4-88.1 |
| Yes | 6,302 | 12.2 | 11.9-12.6 |
| **Hearing related disabilities** |  |  |  |
| No | 50,286 | 97.6 | 97.4-97.7 |
| Yes | 1,249 | 2.4 | 2.3-2.6 |
| **Walking disabilities** |  |  |  |
| No | 47,310 | 91.8 | 91.5-92.1 |
| Yes | 4,225 | 8.2 | 7.9-8.5 |
| **Cognitive disabilities** |  |  |  |
| No | 46,645 | 90.5 | 90.2-90.8 |
| Yes | 4,890 | 9.5 | 9.2-9.8 |
| **Self-care related disabilities** |  |  |  |
| No | 50,964 | 98.9 | 98.8-99.0 |
| Yes | 571 | 1.1 | 1.0-1.2 |
| **Communication related disabilities** |  |  |  |
| No | 51,190 | 99.3 | 99.3-99.4 |
| Yes | 344 | 0.7 | 0.6-0.8 |

### Distribution of menstrual hygiene management practice and its consequences in general and among women with disability

We presented the percentages of MHM, both in general and among women with disabilities, in Table 3. Approximately 4% of the total respondents reported inappropriate use of materials while menstruating to manage blood flow, followed by around 3% reporting having no private place for washing and changing menstrual materials. About 7.5% of the total sample reported that their social activities were impacted because of menstruation. These percentages increased for women with disabilities, with higher percentages observed among those with severe disabilities followed by moderate disabilities and no disabilities. A total of 8% of women with severe disabilities reported inappropriate use of materials to manage blood flow, followed by 6.5% reporting having no private place to wash menstrual materials, and 11.0% reporting that their social lives were impacted because of menstruation.

**Table 3:** Menstrual hygiene management practice in general and across persons with disability.

| Outcomes | Overall percentage<br>(95% CI) | Disability Status, % (95% CI) |  |  | P-value+ |
| --- | --- | --- | --- | --- | --- |
|  |  | Women with<br>severe disabilities | Women with<br>moderate<br>disabilities | Women with no<br>disabilities |  |
| **Material used to manage blood flow while menstruating** |  |  |  |  |  |
| Not appropriate | 3.9 (3.7-4.1) | 8.0 (6.5-9.7) | 5.8 (5.3-6.3) | 3.3 (3.0-3.5) | <0.01 |
| Appropriate | 96.1 (95.9-96.3) | 92.0 (90.3-93.5) | 94.2 (93.7-94.7) | 96.8 (96.5-97.0) |  |
| **Availability of private place for washing and changing while at home** |  |  |  |  |  |
| No | 3.2 (3.0-3.4) | 6.5 (5.1-8.3) | 4.3 (3.9-4.8) | 2.9 (2.6-3.1) | <0.01 |
| Yes | 96.8 (96.6-97.0) | 93.5 (91.8-94.9) | 95.7 (95.2-96.1) | 97.2 (96.9-97.4) |  |
| **Menstruation impacted in participation of social activities, including schooling and work** |  |  |  |  |  |
| Not impacted | 92.5 (92.2-92.7) | 89.0 (87.3-90.6) | 93.1 (92.5-93.6) | 92.4 (92.1-92.7) | <0.01 |
| Impacted | 7.5 (7.3-7.8) | 11.0 (9.5-12.7) | 6.9 (6.4-7.5) | 7.6 (7.3-7.9) |  |

Note: \*p-values were obtained from chi-square test in accessing differences between menstrual hygiene management practice and consequences across level of disabilities

### Distribution of level of disabilities across selected covariates

The distribution of disabilities level across considered covariates are presented in S-Table 1. We found highest prevalence of moderate and severe disabilities among women aged 35 and older. Women with no formal education and not exposed to mass media exhibited the higher prevalence of moderate and severe disabilities. In terms of household level factors, among women who resided in the poorest wealth quintile reported the higher prevalence of both sorts of disabilities. Moreover, women who were from Barishal, Khulna, Mymensingh divisions had the highest prevalence of both sorts of disabilities.

**Figure 1.**
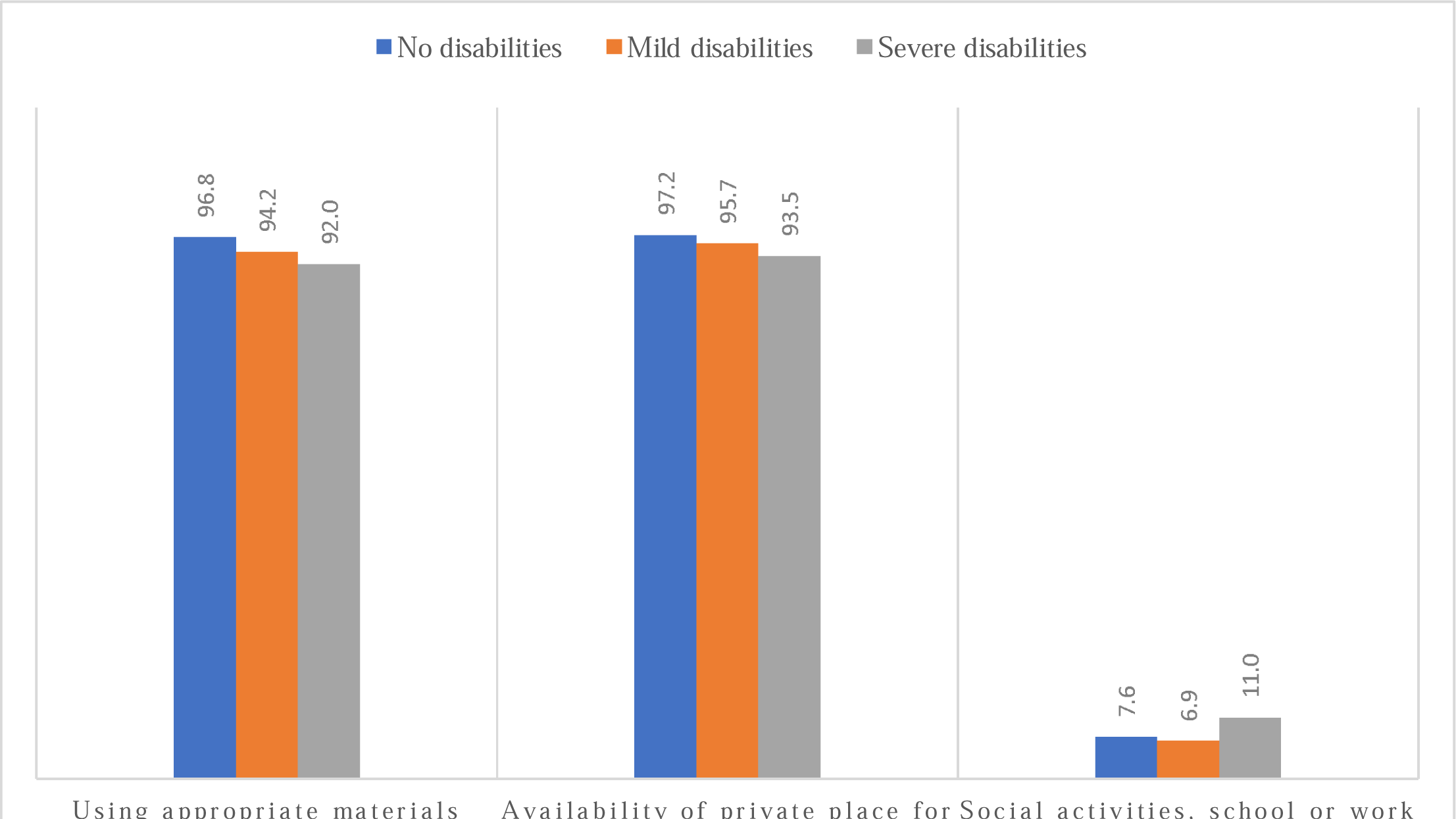
Distribution of menstrual hygiene management across level of disabilities

### Association of level of disabilities with menstrual hygiene management and its consequences

Association between MHM and its consequences with the level of disabilities and disabilities type were assessed both unadjusted and adjusted multilevel mixed effect logistic regression model. The unadjusted model revealed decreased likelihoods of appropriate MHM practices and their consequences with increasing severity and types of disabilities (S-Table 2). These findings remained consistent after adjusting for socio-demographic factors of the respondents (Table 4 and S-Table 3). We found 33-47% lower likelihoods of using appropriate materials to manage menstrual flow among women with higher levels of disabilities compared to those without disabilities. Similarly, women with moderate to severe level of disabilities reported 34-54% lower likelihoods of having access to private spaces at home for washing and changing menstrual rags, compared to women without disabilities. Moreover, women with severe disabilities were 1.66 times more likely (95% CI, 1.35-2.03) to report that their participation in social activities, school, or work was impacted during menstruation, compared to women without disabilities. Among the adjusted factors, older age was associated with inappropriate MHM practices, lack of private spaces at home for changing menstrual rags, and reduced attendance in social activities. Additionally, women with higher education levels and those residing in wealthier households, urban areas, and the Sylhet division reported higher likelihoods of practicing positive MHM behaviors. These reported findings were found consistent once we segregate these results across types of disabilities (S-Table 4-6).

**Table 4.** Adjusted associations from multilevel mixed-effects logistic regression model of assessing associations of menstrual hygiene management and its consequences with disabilities label and type of disabilities adjusted for individual, household, and community level factors in Bangladesh, 2019 (N= 51,535)

| Exposures | Appropriate material used to manage blood flow while menstruating<br>aOR (95% CI)+ | Availability of a private place at home for washing and changing menstruation rags (yes, no)<br>aOR (95% CI) | Impacted in attendance of social activities, school, or work during menstruation<br>aOR (95% CI) |
| --- | --- | --- | --- |
| **Disability level** |  |  |  |
| No disabilities (ref) | 1.00 | 1.00 | 1.00 |
| Moderate disabilities | 0.67 (0.60-0.75)*** | 0.66 (0.58-0.76)*** | 0.91 (0.82-1.01) |
| Severe disabilities | 0.53 (0.42-0.67)*** | 0.46 (0.35-0.59)*** | 1.66 (1.35-2.03)*** |
| **Domains of disabilities** |  |  |  |
| Vision related disabilities | 0.66 (0.58-0.75)*** | 0.80 (0.69-0.94)*** | 0.83 (0.74-0.94)*** |
| Hearing related disabilities | 0.78 (0.61-0.98)** | 0.51 (0.39-0.65)*** | 1.36 (1.09-1.70)*** |
| Walking disabilities | 0.71 (0.61-0.82)*** | 0.80 (0.67-0.96)** | 0.76 (0.65-0.88)*** |
| Cognitive disabilities | 0.82 (0.70-0.95)*** | 0.47 (0.40-0.54)*** | 1.26 (1.12-1.43)*** |
| Self-care related disabilities | 0.71 (0.50-1.02) | 0.53 (0.36-0.77)*** | 0.97 (0.68-1.38) |
| Communication related disabilities | 0.93 (0.55-1.56) | 0.36 (0.23-0.57)*** | 1.20 (0.80-1.82) |

**Notes:** Models were adjusted with women's age, education, mass media exposure, wealth index, place of residence, and divisions. aOR: Adjusted
Odds Ratios. 95% CI: 95% Confidence Intervals. \*\*\*p<0.01, \*\*p<0.05.

## Discussion

The aim of this study was to explore the effects of disability on MHM and its associated adverse consequences in Bangladesh. Approximately 22% of our analyzed sample had a disability, with 2.7% classified as severe disability and 19.2% as moderate disability. Notably, 3.9% of the overall sample reported using inappropriate materials to manage menstrual flow. This issue is even more prevalent among women with disabilities, reaching 8.0% for those with severe disabilities and 5.8% for those with moderate disabilities. Furthermore, 6.5% of women with severe disabilities reported not having a private place for washing and changing menstrual rags at home, while 11% reported their social activities being impacted by menstruation. After adjusting for socio-demographic factors, we found significantly lower odds (33-47%) of not using appropriate materials to manage menstrual flow among women with moderate to severe disabilities compared to those without disabilities. Similarly, the likelihood of having a private place to change menstrual rags at home was 34-44% lower among women with moderate to severe disabilities. Additionally, women with severe disabilities had a 1.66 times higher likelihood of reporting that their social activities were impacted by menstruation compared to women without disabilities. These findings are robust, being derived from a large-scale nationally representative survey and adjusting for a broad range of socio-demographic factors. This study underscores the significant challenges faced by women with disabilities in managing menstrual hygiene and highlights the importance of tailored interventions to address their specific needs.

The findings indicate a significant disparity in menstrual hygiene management among women with disabilities compared to those without. Specifically, women with severe disabilities were about three times more likely, and those with moderate disabilities were around twice as likely, to report not using appropriate menstrual products. These observations are consistent with available literature in LMICs and Bangladesh [30, 43, 44]. This suggests multiple potential underlying issues. Accessibility and availability are critical as women with disabilities may face difficulties accessing menstrual products due to physical barriers, lack of transportation, or unavailability of products in nearby locations [36, 37, 45]. Economic factors further complicate this issue, as disabilities can impact economic stability, making it harder for women to afford menstrual products [46]. Education and awareness are often lacking among women with disabilities, with societal neglect in providing inclusive health education [47]. Furthermore, societal stigma and misconceptions surrounding disability and menstruation may contribute to a lack of awareness and education on proper menstrual hygiene practices among this population group [7, 48]. Stigma and discrimination around menstruation and disabilities also result in inadequate support from families and communities, limiting effective menstrual health management. Additionally, healthcare systems might not be adequately equipped or sensitive to the needs of women with disabilities, leading to insufficient guidance and support [49, 50]. Women with disabilities are also more likely to report a lack of private spaces at home where they can wash and change menstrual rags. This observation is closely reflected in Bangladeshi culture and is comparable to findings from previous studies in Bangladesh and other LMICs [36, 37, 51]. In a typical Bangladeshi household, family members usually share a common toilet, which women also use for changing menstrual rags and washing them, with no separate space dedicated to these activities [52]. While non-disabled women can manage by washing menstrual rags in the toilet and drying them in hidden areas within the household, women with disabilities face additional challenges due to restricted mobility. This issue is particularly challenging for women with disabilities who need assistance to go to the toilet, as they lack the privacy and autonomy to manage their menstrual hygiene independently [36]. The situation is exacerbated by limited access to accessible sanitation facilities and the cultural stigma surrounding menstruation, which further restricts the options available to these women [35, 37].

This study found that women with disabilities are more likely to report that their social activities are impacted by their menstrual periods compared to women without disabilities. This broader impact can be attributed to several factors. Firstly, women with disabilities often face mobility challenges, which can make it difficult to manage menstrual hygiene effectively. Secondly, the lack of accessible and private facilities for changing menstrual products exacerbates the problem, leading to greater social isolation during menstruation. Additionally, societal stigma and lack of awareness about the needs of women with disabilities can contribute to their social exclusion. In many LMICs, including Bangladesh, these issues are intensified by inadequate infrastructure and support systems [2, 27, 36, 37].

The findings of this study have several policy implications. Poor MHM among women with disabilities indicates the need for targeted interventions to improve accessibility and availability of menstrual hygiene products and facilities. Policies should focus on providing accessible sanitation infrastructure, ensuring economic support for women with disabilities, and promoting inclusive health education that addresses menstrual hygiene management. Raising awareness and reducing stigma around menstruation and disabilities within communities is crucial. Additionally, healthcare systems should be equipped to support the specific needs of women with disabilities, ensuring they receive adequate guidance and assistance for MHM.

This study possesses several strengths alongside a few limitations. One of its primary strengths is the extensive reach of the MICS, facilitating the inclusion of diverse geographic regions and socio-demographic groups, thereby enhancing the representativeness of findings. Furthermore, the cross-sectional nature of the survey enables the collection of data from a large sample within a relatively short timeframe, providing researchers with a snapshot of MHM practices and challenges across the country. The data analysis was conducted comprehensively using sophisticated statistical methods, considering a range of socio-demographic characteristics of the respondents. This comprehensive approach offers insights into the prevalence of various MHM practices among women with disabilities, illuminating both common trends and disparities across different population groups. However, the cross-sectional design of this study inherently limits the ability to establish causal relationships or track changes over time. Additionally, reliance on self-reported data may introduce bias, as participants may underreport or overreport certain behaviors or experiences related to menstrual hygiene due to social desirability or recall bias. Moreover, while the MICS aims to be nationally representative, marginalized groups or individuals with disabilities may still be underrepresented or excluded from the survey sample, potentially constraining the generalizability of findings to the entire population of women with disabilities in Bangladesh. Furthermore, despite considering various factors in the model, social norms and stigma affecting MHM were not incorporated due to a lack of relevant data in the survey. These limitations underscore the necessity for complementary qualitative research and targeted interventions to augment insights gained from the MICS data, ensuring a more nuanced understanding of the intricate interplay between disabilities and MHM practices in Bangladesh.

## Conclusion

The findings indicate that women with disabilities, particularly those with severe disabilities, are more likely to encounter obstacles in accessing appropriate menstrual products and private spaces for hygiene management. Additionally, their social activities are disproportionately affected by menstruation, highlighting the multifaceted impact of disabilities on menstrual health. These findings underscore the urgent need for tailored interventions to address the specific needs of women with disabilities in Bangladesh. Accessibility and availability of menstrual hygiene products and facilities must be improved, alongside efforts to enhance economic support and inclusive health education. Raising awareness and reducing stigma surrounding menstruation and disabilities within communities is essential to promote social inclusion and support networks for women with disabilities.

## Supporting information

https://mics.unicef.org/surveys

## Data Availability

The datasets used and analyzed in this study are available from the UNICEF MICS Archive website: https://mics.unicef.org/surveys

## Declarations

## Acknowledgement

We would like to express our gratitude to the UNICEF MICS for granting access to the 2019 BMICS data to conduct the study. Moreover, we also acknowledge the support of Department of Population science, Jatiya Kabi Kazi Nazrul Islam University, Bangladesh, where the study was conducted.

## Authors’ contributors

MNK and MBA designed the study concept. MBA conducted the formal analysis with the help of MNK. MNK drafted the manuscript. MNK, SJK, RH and MAK reviewed the first manuscript. MNK supervised all works. All authors critically reviewed and approved the final version of this manuscript.

