## Supplementary material for "Menstrual hygiene management among reproductive-aged women with disabilities in Bangladesh": https://mics.unicef.org/surveys

**S-Table 1. Level of disabilities by respondent’s background characteristics**

| **Characteristics** | **No disabilities, % (95% CI)** | **Moderate disabilities, % (95% CI)** | **Severe disabilities, % (95% CI)** |
| --- | --- | --- | --- |
| **Individual level factors** |  |  |  |
| **Ages of women** |  |  |  |
| ≤24 years | 90.5 (90.0-91.1) | 8.5 (8.0-9.1) | 1.0 (0.8-1.2) |
| 25-34 years | 83.2 (82.5-83.8) | 15.0 (14.4-15.7) | 1.8 (1.6-2.0) |
| ≥35 years | 62.7 (61.9-63.6) | 32.3 (31.5-33.1) | 5.0 (4.6-5.3) |
| **Women’s education** |  |  |  |
| Pre-primary | 67.6 (66.4-68.8) | 27.8 (26.6-28.9) | 4.8 (4.2-5.2) |
| Primary | 73.6 (72.7-74.5) | 22.9 (22.0-23.8) | 3.5 (3.2-3.9) |
| Secondary | 81.5 (80.8-82.1) | 16.5 (15.9-17.1) | 2.1 (1.9-2.3) |
| Higher | 85.6 (84.6-86.5) | 13.2 (12.4-14.1) | 1.2 (1.0-1.5) |
| **Exposure to mass media** |  |  |  |
| Not exposed | 74.3 (73.5-75.1) | 21.9 (21.2-22.7) | 3.8 (3.5-4.1) |
| Exposed | 79.8 (79.3-80.4) | 18.0 (17.5-18.5) | 2.2 (2.0-2.4) |
| **Household level factors** |  |  |  |
| **Wealth index** |  |  |  |
| Poorest | 74.7 (73.6-75.8) | 21.4 (20.4-22.4) | 3.9 (3.5-4.4) |
| Second | 77.8 (76.8-78.7) | 19.6 (18.7-20.5) | 2.6 (2.3-3.0) |
| Middle | 78.6 (77.6-79.5) | 19.0 (18.1-19.9) | 2.5 (2.2-2.8) |
| Fourth | 80.1 (79.1-81.0) | 17.7 (16.8-18.6) | 2.3 (2.0-2.6) |
| Richest | 79.1 (77.9-80.1) | 18.7 (17.7-19.7) | 2.3 (2.0-2.7) |
| **Community level factors** |  |  |  |
| **Place of residence** |  |  |  |
| Urban | 77.7 (76.5-78.8) | 19.6 (18.6-20.7) | 2.7 (2.4-3.1) |
| Rural | 78.3 (77.8-78.8) | 19.0 (18.6-19.5) | 2.7 (2.5-2.8) |
| **Divisions** |  |  |  |
| Barishal | 63.1 (61.5-64.7) | 29.3 (27.8-30.8) | 7.7 (6.8-8.6) |
| Chattogram | 80.4 (79.3-81.3) | 16.5 (15.6-17.4) | 3.2 (2.8-3.6) |
| Dhaka | 82.8 (81.7-83.9) | 15.6 (14.6-16.6) | 1.6 (1.4-1.9) |
| Khulna | 67.6 (66.4-68.8) | 28.5 (27.4-29.7) | 3.9 (3.4-4.5) |
| Mymensingh | 73.5 (71.6-75.3) | 22.7 (20.9-24.5) | 3.9 (3.1-4.7) |
| Rajshahi | 77.7 (76.3-78.9) | 20.6 (19.4-21.8) | 1.8 (1.4-2.2) |
| Rangpur | 84.8 (83.6-85.9) | 14.0 (12.9-15.1) | 1.2 (1.0-1.6) |
| Sylhet | 79.0 (77.3-80.6) | 18.8 (17.2-20.6) | 2.2 (1.7-2.8) |

**Note:** Presented as row percentages.

**S-Table 2.** Unadjusted association from multilevel mixed-effects logistic regression model of assessing associations of menstrual hygiene management and its consequences with disabilities label and type of disabilities adjusted for individual, household, and community level factors in Bangladesh, 2019 **(N= 51,535)**

| **Exposures** | **Appropriate material used to manage blood flow while menstruating** | **Availability of private place for washing and changing while at home** | **Menstruation impacted in participation of social activities, including schooling and work** |
| --- | --- | --- | --- |
|  | **cOR (95% CI)** | **cOR (95% CI)** | **cOR (95% CI)** |
| **Disability level** |  |  |  |
| No disabilities (ref) | 1.00 | 1.00 | 1.00 |
| Moderate disabilities | 0.49 (0.44-0.55)** | 0.66 (0.58-0.75)*** | 0.86 (0.78-0.95)*** |
| Severe disabilities | 0.35 (0.28-0.44)** | 0.43 (0.33-0.54)*** | 1.61 (1.32-1.97)*** |
| **Domains of disabilities** |  |  |  |
| Vision | 0.43 (0.38-0.49)** | 0.76 (0.65-0.88)*** | 0.81 (0.72-0.91)*** |
| Hearing | 0.57 (0.44-0.72)** | 0.48 (0.37-0.62)*** | 1.29 (1.03-1.61)** |
| Mobility | 0.53 (0.46-0.62)** | 0.80 (0.67-0.95)** | 0.72 (0.62-0.83)*** |
| Cognition | 0.61 (0.53-0.71)** | 0.45 (0.39-0.52)*** | 1.22 (1.08-1.38)*** |
| Self-care related disabilities | 0.47 (0.33-0.67)** | 0.48 (0.33-0.71)*** | 0.58 (0.47-0.73)*** |
| Communication disabilities | 0.69 (0.42-1.15) | 0.33 (0.21-0.51)*** | 1.22 (0.81-1.85) |

**Notes:** cOR: Crude Odds Ratios. 95% CI: 95% Confidence Intervals. ***p<0.01, **p<0.05.

**S-Table 3:** Adjusted association from from multilevel mixed-effects logistic regression model of assessing associations of menstrual hygiene management and its consequences with disabilities label and type of disabilities adjusted for individual, household, and community level factors in Bangladesh, 2019 **(N= 51,535)**

| **Exposures** | **Appropriate material used to manage blood flow while menstruating** | **Availability of a private place at home for washing and changing menstruation rags (yes, no)** | **Impacted in attendance of social activities, school, or work during menstruation** |
| --- | --- | --- | --- |
|  | **aOR (95% CI)+** | **aOR (95% CI)** | **aOR (95% CI)** |
| **Disability level** |  |  |  |
| No disabilities (ref) | 1.00 | 1.00 | 1.00 |
| Moderate disabilities | 0.67 (0.60-0.75)*** | 0.66 (0.58-0.76)*** | 0.91 (0.82-1.01) |
| Severe disabilities | 0.53 (0.42-0.67)*** | 0.46 (0.35-0.59)*** | 1.66 (1.35-2.03)*** |
| **Individual level factors** |  |  |  |
| **Ages of women** |  |  |  |
| ≤24 years | 1.00 | 1.00 | 1.00 |
| 25-34 years | 0.53 (0.45-0.62)*** | 1.03 (0.90-1.18) | 0.90 (0.83-0.99)** |
| ≥35 years | 0.29 (0.24-0.34)*** | 0.97 (0.83-1.13) | 0.82 (0.74-0.91)*** |
| **Women’s education** |  |  |  |
| Pre-primary | 1.00 | 1.00 | 1.00 |
| Primary | 1.15 (1.02-1.31)** | 0.93 (0.79-1.09) | 0.82 (0.73-0.92)*** |
| Secondary | 1.54 (1.34-1.77)*** | 1.07 (0.91-1.27) | 0.86 (0.76-0.97)** |
| Higher | 2.92 (2.30-3.71)*** | 1.00 (0.81-1.25) | 0.91 (0.78-1.06) |
| **Exposure to mass media** |  |  |  |
| Not exposed | 1.00 | 1.00 | 1.00 |
| Exposed | 1.02 (0.92-1.15) | 1.08 (0.95-1.23) | 0.99 (0.91-1.09) |
| **Household level factors** |  |  |  |
| **Wealth index** |  |  |  |
| Poorest | 1.00 | 1.00 | 1.00 |
| Second | 1.22 (1.05-1.42)*** | 1.28 (1.09-1.52)*** | 1.00 (0.88-1.15) |
| Middle | 1.28 (1.09-1.50)*** | 1.39 (1.16-1.66)*** | 0.95 (0.83-1.09) |
| Fourth | 1.23 (1.04-1.46)*** | 1.30 (1.07-1.58)*** | 0.89 (0.78-1.03) |
| Richest | 1.91 (1.52-2.39)*** | 1.57 (1.24-1.99)*** | 0.84 (0.71-0.98)** |
| **Community level factors** |  |  |  |
| **Place of residence** |  |  |  |
| Urban | 1.00 | 1.00 | 1.00 |
| Rural | 0.74 (0.62-0.89)*** | 1.09 (0.89-1.34) | 1.29 (1.08-1.54)*** |
| **Divisions** |  |  |  |
| Barishal | 1.00 | 1.00 | 1.00 |
| Chattogram | 0.60 (0.46-0.78)*** | 0.94 (0.69-1.28) | 1.19 (0.91-1.55) |
| Dhaka | 0.85 (0.65-1.11) | 2.15 (1.56-2.97)*** | 0.80 (0.61-1.04) |
| Khulna | 1.95 (1.43-2.68)*** | 1.82 (1.29-2.57)*** | 0.51 (0.38-0.68)*** |
| Mymensingh | 0.76 (0.56-1.04) | 0.57 (0.40-0.82)*** | 0.12 (0.08-0.19)*** |
| Rajshahi | 2.15 (1.57-2.95)*** | 2.41 (1.69-3.45)*** | 0.28 (0.20-0.38)*** |
| Rangpur | 0.52 (0.39-0.69)*** | 0.48 (0.35-0.66)*** | 0.25 (0.18-0.35)*** |
| Sylhet | 7.98 (4.65-13.67)*** | 2.76 (1.78-4.29)*** | 2.37 (1.73-3.24)*** |

**S-Table 4.** Results from multilevel mixed-effects logistic regression model of appropriate material used to manage blood flow while menstruating among women with several domains of disabilities adjusted with individual, household, and community level factors in Bangladesh, 2019

| **Exposures** | **Vision related disabilities** | **Hearing related disabilities** | **Walking disabilities** | **Cognitive disabilities** | **Self-care related disabilities** | **Communication related disabilities** |
| --- | --- | --- | --- | --- | --- | --- |
|  | **aOR (95% CI)** | **aOR (95% CI)** | **aOR (95% CI)** | **aOR (95% CI)** | **aOR (95% CI)** | **aOR (95% CI)** |
| **Material used to manage blood flow while menstruating** |  |  |  |  |  |  |
| Not-appropriate | 1.00 | 1.00 | 1.00 | 1.00 | 1.00 | 1.00 |
| Appropriate | 0.66 (0.58-0.75)*** | 0.78 (0.61-0.98)** | 0.71 (0.61-0.82)*** | 0.82 (0.70-0.95)*** | 0.71 (0.50-1.02) | 0.93 (0.55-1.56) |
| **Individual level factors** |  |  |  |  |  |  |
| **Ages of women** |  |  |  |  |  |  |
| ≤24 years | 1.00 | 1.00 | 1.00 | 1.00 | 1.00 | 1.00 |
| 25-34 years | 0.52 (0.44-0.61)*** | 0.51 (0.43-0.61)*** | 0.52 (0.44-0.61)*** | 0.51 (0.44-0.61)*** | 0.51 (0.44-0.61)*** | 0.51 (0.44-0.60)*** |
| ≥35 years | 0.28 (0.24-0.33)*** | 0.26 (0.22-0.31)*** | 0.27 (0.23-0.32)*** | 0.26 (0.22-0.31)*** | 0.26 (0.22-0.31)*** | 0.26 (0.22-0.31)*** |
| **Women’s education** |  |  |  |  |  |  |
| Pre-primary | 1.00 | 1.00 | 1.00 | 1.00 | 1.00 | 1.00 |
| Primary | 1.15 (1.02-1.31)** | 1.16 (1.02-1.32)*** | 1.16 (1.02-1.31)** | 1.16 (1.02-1.32)** | 1.16 (1.02-1.32)** | 1.16 (1.02-1.32)** |
| Secondary | 1.54 (1.34-1.77)*** | 1.57 (1.36-1.80)*** | 1.56 (1.36-1.79)*** | 1.56 (1.36-1.80)*** | 1.57 (1.36-1.80)*** | 1.57 (1.36-1.80)*** |
| Higher | 2.96 (2.33-3.75)*** | 3.02 (2.38-3.83)*** | 2.99 (2.36-3.79)*** | 3.01 (2.37-3.52)*** | 3.02 (2.38-3.83)*** | 3.04 (2.39-3.85)*** |
| **Exposure to mass media** |  |  |  |  |  |  |
| Not exposed | 1.00 | 1.00 | 1.00 | 1.00 | 1.00 | 1.00 |
| Exposed | 1.03 (0.92-1.16) | 1.04 (0.93-1.17) | 1.04 (0.92-1.16) | 1.04 (0.92-1.16) | 1.04 (0.92-1.16) | 1.04 (0.93-1.17) |
| **Household level factors** |  |  |  |  |  |  |
| **Wealth index** |  |  |  |  |  |  |
| Poorest | 1.00 | 1.00 | 1.00 | 1.00 | 1.00 | 1.00 |
| Second | 1.22 (1.06-1.42)*** | 1.21 (1.05-1.41)** | 1.22 (1.06-1.42)*** | 1.21 (1.05-1.41)** | 1.22 (1.05-1.41)** | 1.22 (1.05-1.41)** |
| Middle | 1.28 (1.09-1.49)*** | 1.26 (1.08-1.48)*** | 1.28 (1.09-1.50)*** | 1.26 (1.08-1.48)*** | 1.27 (1.08-1.48)*** | 1.27 (1.08-1.48)*** |
| Fourth | 1.23 (1.03-1.46)** | 1.21 (1.02-1.44)*** | 1.23 (1.03-1.46)** | 1.21 (1.02-1.44)** | 1.22 (1.02-1.45)** | 1.21 (1.02-1.44)** |
| Richest | 1.89 (1.51-2.37)*** | 1.85 (1.48-2.32)*** | 1.90 (1.51-2.38)*** | 1.86 (1.49-2.33)*** | 1.86 (1.48-2.33)*** | 1.86 (1.48-2.32)*** |
| **Community level factors** |  |  |  |  |  |  |
| **Place of residence** |  |  |  |  |  |  |
| Urban | 1.00 | 1.00 | 1.00 | 1.00 | 1.00 | 1.00 |
| Rural | 0.75 (0.63-0.89)*** | 0.75 (0.62-0.89)*** | 0.74 (0.62-0.89)*** | 0.74 (0.62-0.89)*** | 0.74 (0.62-0.89)*** | 0.75 (0.62-0.89)*** |
| **Divisions** |  |  |  |  |  |  |
| Barishal | 1.00 | 1.00 | 1.00 | 1.00 | 1.00 | 1.00 |
| Chattogram | 0.63 (0.48-0.82)*** | 0.65 (0.50-0.85)*** | 0.64 (0.49-0.84)*** | 0.64 (0.49-0.84)*** | 0.66 (0.50-0.86)*** | 0.66 (0.51-0.86)*** |
| Dhaka | 0.91 (0.70-1.20) | 0.95 (0.73-1.24) | 0.92 (0.70-1.20) | 0.92 (0.70-1.20) | 0.95 (0.72-1.24) | 0.96 (0.73-1.25) |
| Khulna | 2.00 (1.46-2.74)*** | 1.99 (1.46-2.73)*** | 2.02 (1.48-2.77)*** | 1.98 (1.44-2.71)*** | 1.99 (1.45-2.72)*** | 2.00 (1.46-2.73)*** |
| Mymensingh | 0.79 (0.58-1.08) | 0.81 (0.59-1.10) | 0.79 (0.58-1.09) | 0.79 (0.58-1.09) | 0.80 (0.59-1.10) | 0.81 (0.59-1.11) |
| Rajshahi | 2.28 (1.66-3.12)*** | 2.33 (1.70-3.19)*** | 2.30 (1.68-3.14)*** | 2.26 (1.65-3.10)*** | 2.32 (1.69-3.17)*** | 2.34 (1.71-3.20)*** |
| Rangpur | 0.56 (0.43-0.74)*** | 0.59 (0.50-0.78)*** | 0.57 (0.43-0.75)*** | 0.57 (0.43-0.75)*** | 0.59 (0.45-0.77)*** | 0.60 (0.45-0.78)*** |
| Sylhet | 8.45 (4.94-14.48)*** | 8.61 (5.03-14.73)*** | 8.52 (4.97-14.58)*** | 8.44 (4.93-14.46)*** | 8.60 (5.03-14.73)*** | 8.65 (5.06-14.81)*** |

**S-Table 5.** Multilevel mixed-effects logistic regression model of availability of private place for washing and changing while at home among menstruated women with several domains of disabilities adjusted with individual, household, and community level factors in Bangladesh, 2019

| **Exposures** | **Vision related disabilities** | **Hearing related disabilities** | **Walking disabilities** | **Cognitive disabilities** | **Self-care related disabilities** | **Communication related disabilities** |
| --- | --- | --- | --- | --- | --- | --- |
|  | **aOR (95% CI)** | **aOR (95% CI)** | **aOR (95% CI)** | **aOR (95% CI)** | **aOR (95% CI)** | **aOR (95% CI)** |
| **Availability of private place for washing and changing while at home** |  |  |  |  |  |  |
| No | 1.00 | 1.00 | 1.00 | 1.00 | 1.00 | 1.00 |
| Yes | 0.80 (0.69-0.94)*** | 0.51 (0.39-0.65)*** | 0.80 (0.67-0.96)** | 0.47 (0.40-0.54)*** | 0.53 (0.36-0.77)*** | 0.36 (0.23-0.57)*** |
| **Individual level factors** |  |  |  |  |  |  |
| **Ages of women** |  |  |  |  |  |  |
| ≤24 years | 1.00 | 1.00 | 1.00 | 1.00 | 1.00 | 1.00 |
| 25-34 years | 1.00 (0.87-1.15) | 1.01 (0.88-1.16) | 1.01 (0.88-1.15) | 1.02 (0.89-1.17) | 1.00 (0.87-1.15) | 1.00 (0.87-1.14) |
| ≥35 years | 0.90 (0.77-1.04) | 0.88 (0.76-1.02) | 0.89 (0.76-1.03) | 0.93 (0.80-1.08) | 0.87 (0.75-1.01) | 0.86 (0.74-1.00) |
| **Women’s education** |  |  |  |  |  |  |
| Pre-primary | 1.00 | 1.00 | 1.00 | 1.00 | 1.00 | 1.00 |
| Primary | 0.93 (0.79-1.10) | 0.93 (0.79-1.09) | 0.93 (0.79-1.09) | 0.92 (0.78-1.09) | 0.93 (0.79-1.15) | 0.93 (0.79-1.10) |
| Secondary | 1.09 (0.92-1.29) | 1.09 (0.92-1.29) | 1.10 (0.93-1.30) | 1.07 (0.90-1.27) | 1.10 (0.92-1.30) | 1.09 (0.92-1.29) |
| Higher | 1.04 (0.83-1.29) | 1.03 (0.83-1.28) | 1.04 (0.84-1.29) | 1.00 (0.80-1.24) | 1.04 (0.84-1.29) | 1.03 (0.83-1.28) |
| **Exposure to mass media** |  |  |  |  |  |  |
| Not exposed | 1.00 | 1.00 | 1.00 | 1.00 | 1.00 | 1.00 |
| Exposed | 1.09 (0.95-1.24) | 1.09 (0.96-1.24) | 1.09 (0.96-1.24) | 1.07 (0.94-1.22) | 1.09 (0.96-1.24) | 1.09 (0.95-1.24) |
| **Household level factors** |  |  |  |  |  |  |
| **Wealth index** |  |  |  |  |  |  |
| Poorest | 1.00 | 1.00 | 1.00 | 1.00 | 1.00 | 1.00 |
| Second | 1.30 (1.10-1.53)*** | 1.29 (1.09-1.52)*** | 1.30 (1.10-1.53)*** | 1.28 (1.08-1.51)*** | 1.30 (1.10-1.53)*** | 1.30 (1.10-1.53)*** |
| Middle | 1.40 (1.17-1.67)*** | 1.39 (1.16-1.66)*** | 1.40 (1.17-1.68)*** | 1.38 (1.16-1.64)*** | 1.40 (1.17-1.68)*** | 1.39 (1.17-1.67)*** |
| Fourth | 1.31 (1.08-1.58)*** | 1.29 (1.07-1.57)*** | 1.31 (1.08-1.59)*** | 1.29 (1.07-1.57)*** | 1.31 (1.08-1.59)*** | 1.31 (1.08-1.59)*** |
| Richest | 1.57 (1.24-1.99)*** | 1.55 (1.22-1.96)*** | 1.58 (1.24-2.00)*** | 1.55 (1.22-1.97)*** | 1.57 (1.24-1.98)*** | 1.57 (1.24-1.98)*** |
| **Community level factors** |  |  |  |  |  |  |
| **Place of residence** |  |  |  |  |  |  |
| Urban | 1.00 | 1.00 | 1.00 | 1.00 | 1.00 | 1.00 |
| Rural | 1.11 (0.91-1.36) | 1.11 (0.91-1.36) | 1.11 (0.90-1.35) | 1.09 (0.89-1.33) | 1.10 (0.90-1.35) | 1.10 (0.90-1.35) |
| **Divisions** |  |  |  |  |  |  |
| Barishal | 1.00 | 1.00 | 1.00 | 1.00 | 1.00 | 1.00 |
| Chattogram | 1.01 (0.74-1.38) | 1.02 (0.75-1.38) | 1.02 (0.75-1.39) | 0.92 (0.68-1.25) | 1.03 (0.76-1.40) | 1.04 (0.76-1.42) |
| Dhaka | 2.36 (1.71-3.26)*** | 2.38 (1.72-3.28)*** | 2.36 (1.71-3.25)*** | 2.09 (1.51-2.88)*** | 2.36 (1.71-3.26)*** | 2.40 (1.74-3.32)*** |
| Khulna | 1.87 (1.32-2.64)*** | 1.88 (1.33-2.66)*** | 1.89 (1.34-2.67)*** | 1.80 (1.28-2.53)*** | 1.86 (1.31-2.62)*** | 1.90 (1.34-2.68)*** |
| Mymensingh | 0.60 (0.42-0.86)*** | 0.61 (0.43-0.87)*** | 0.61 (0.45-0.87)*** | 0.58 (0.40-0.82)*** | 0.60 (0.42-0.86)*** | 0.62 (0.43-0.88)*** |
| Rajshahi | 2.60 (1.82-3.72)*** | 2.64 (1.85-3.77)*** | 2.61 (1.83-3.73)*** | 2.33 (1.63-3.33)*** | 2.60 (1.82-3.72)*** | 2.65 (1.85-3.79)*** |
| Rangpur | 0.54 (0.39-0.73)*** | 0.54 (0.80-0.74)*** | 0.54 (0.39-0.74)*** | 0.48 (0.35-0.65)*** | 0.54 (0.40-0.74)*** | 0.55 (0.40-0.75)*** |
| Sylhet | 2.99 (1.92-4.63)*** | 2.99 (1.93-4.64)*** | 3.00 (1.93-4.65)*** | 2.74 (1.77-4.24)*** | 3.00 (1.93-4.65)*** | 3.05 (1.96-4.73)*** |

**S-Table 6.** Multilevel mixed-effects logistic regression model of availability of menstruation impacted in participation of social activities, including schooling and work among women with several domains of disabilities adjusted with individual, household, and community level factors in Bangladesh, 2019

| **Exposures** | **Vision related disabilities** | **Hearing related disabilities** | **Walking disabilities** | **Cognitive disabilities** | **Self-care related disabilities** | **Communication related disabilities** |
| --- | --- | --- | --- | --- | --- | --- |
|  | **aOR (95% CI)** | **aOR (95% CI)** | **aOR (95% CI)** | **aOR (95% CI)** | **aOR (95% CI)** | **aOR (95% CI)** |
| **Availability of private place for washing and changing while at home** |  |  |  |  |  |  |
| No | 1.00 | 1.00 | 1.00 | 1.00 | 1.00 | 1.00 |
| Yes | 0.83 (0.74-0.94)*** | 1.36 (1.09-1.70)*** | 0.76 (0.65-0.88)*** | 1.26 (1.12-1.43)*** | 0.97 (0.68-1.38) | 1.20 (0.80-1.82) |
| **Individual level factors** |  |  |  |  |  |  |
| **Ages of women** |  |  |  |  |  |  |
| ≤24 years | 1.00 | 1.00 | 1.00 | 1.00 | 1.00 | 1.00 |
| 25-34 years | 0.91 (0.83-0.99)** | 0.90 (0.82-0.99)** | 0.91 (0.83-1.00) | 0.90 (0.82-0.98)** | 0.90 (0.83-0.99)** | 0.90 (0.82-0.99)** |
| ≥35 years | 0.85 (0.76-0.94)*** | 0.81 (0.73-0.90)*** | 0.85 (0.76-0.93)*** | 0.80 (0.72-0.89)*** | 0.82 (0.74-0.91)*** | 0.82 (0.74-0.91)*** |
| **Women’s education** |  |  |  |  |  |  |
| Pre-primary | 1.00 | 1.00 | 1.00 | 1.00 | 1.00 | 1.00 |
| Primary | 0.81 (0.72-0.92)*** | 0.82 (0.73-0.92)*** | 0.81 (0.72-0.92)*** | 0.82 (0.73-0.93)*** | 0.82 (0.73-0.92)*** | 0.82 (0.73-0.92)*** |
| Secondary | 0.85 (0.75-0.96)*** | 0.86 (0.77-0.97)** | 0.85 (0.75-0.96)*** | 0.86 (0.77-0.97)** | 0.86 (0.76-0.97)*** | 0.86 (0.76-0.97)*** |
| Higher | 0.90 (0.77-1.04) | 0.91 (0.78-1.06) | 0.90 (0.77-1.04) | 0.92 (0.79-1.07) | 0.91 (0.78-1.05) | 0.91 (0.78-1.06) |
| **Exposure to mass media** |  |  |  |  |  |  |
| Not exposed | 1.00 | 1.00 | 1.00 | 1.00 | 1.00 | 1.00 |
| Exposed | 0.99 (0.90-1.09) | 0.99 (0.90-1.09) | 0.99 (0.90-1.09) | 0.99 (0.91-1.09) | 0.99 (0.90-1.09) | 0.99 (0.90-1.09) |
| **Household level factors** |  |  |  |  |  |  |
| **Wealth index** |  |  |  |  |  |  |
| Poorest | 1.00 | 1.00 | 1.00 | 1.00 | 1.00 | 1.00 |
| Second | 1.00 (0.88-1.14) | 1.00 (0.88-1.14) | 1.00 (0.88-1.14) | 1.00 (0.88-1.14) | 1.00 (0.88-1.14) | 1.00 (0.88-1.14) |
| Middle | 0.95 (0.83-1.08) | 0.95 (0.83-1.08) | 0.95 (0.83-1.09) | 0.95 (0.83-1.09) | 0.95 (0.83-1.08) | 0.95 (0.83-1.08) |
| Fourth | 0.89 (0.77-1.03) | 0.89 (0.77-1.03) | 0.89 (0.77-1.03) | 0.89 (0.77-1.03) | 0.89 (0.77-1.03) | 0.89 (0.77-1.03) |
| Richest | 0.84 (0.71-0.98)** | 0.84 (0.71-0.98)** | 0.84 (0.71-0.99)** | 0.84 (0.71-0.98)** | 0.84 (0.71-0.98)** | 0.83 (0.71-0.98)** |
| **Community level factors** |  |  |  |  |  |  |
| **Place of residence** |  |  |  |  |  |  |
| Urban | 1.00 | 1.00 | 1.00 | 1.00 | 1.00 | 1.00 |
| Rural | 1.29 (1.08-1.53)*** | 1.29 (1.08-1.53)*** | 1.28 (1.07-1.53)*** | 1.29 (1.08-1.53)*** | 1.29 (1.08-1.54)*** | 1.29 (1.08-1.54)*** |
| **Divisions** |  |  |  |  |  |  |
| Barishal | 1.00 | 1.00 | 1.00 | 1.00 | 1.00 | 1.00 |
| Chattogram | 1.16 (0.89-1.52) | 1.17 (0.90-1.53) | 1.16 (0.89-1.52) | 1.21 (0.93-1.58) | 1.17 (0.89-1.53) | 1.17 (0.89-1.53) |
| Dhaka | 0.77 (0.59-1.01) | 0.78 (0.60-1.02) | 0.77 (0.59-1.01) | 0.81 (0.62-1.06) | 0.78 (0.60-1.02) | 0.78 (0.60-1.02) |
| Khulna | 0.50 (0.38-0.67)*** | 0.50 (0.37-0.67)*** | 0.51 (0.38-0.68)*** | 0.51 (0.38-0.68)*** | 0.50 (0.37-0.67)*** | 0.50 (0.37-0.67)*** |
| Mymensingh | 0.12 (0.08-0.19)*** | 0.12 (0.08-0.19)*** | 0.12 (0.08-0.19)*** | 0.12 (0.08-0.19)*** | 0.12 (0.08-0.19)*** | 0.12 (0.08-0.19)*** |
| Rajshahi | 0.27 (0.20-0.37)*** | 0.27 (0.20-0.37)*** | 0.27 (0.20-0.37)*** | 0.28 (0.21-0.38)*** | 0.27 (0.20-0.37)*** | 0.27 (0.20-0.37)*** |
| Rangpur | 0.24 (0.18-0.33)*** | 0.25 (0.18-0.34)*** | 0.24 (0.18-0.33)*** | 0.26 (0.19-0.35)*** | 0.25 (0.18-0.34)*** | 0.25 (0.18-0.34)*** |
| Sylhet | 2.30 (1.68-3.15)*** | 2.32 (1.70-3.18)*** | 2.30 (1.68-3.16)*** | 2.39 (1.74-3.27)*** | 2.31 (1.69-3.17)*** | 2.31 (1.69-3.17)*** |
